# Convergent functional effects of antidepressants in major depressive disorder: a neuroimaging meta-analysis

**DOI:** 10.1101/2023.11.24.23298991

**Authors:** Amin Saberi, Amir Ebneabbasi, Sama Rahimi, Sara Sarebannejad, Zumrut Duygu Sen, Heiko Graf, Martin Walter, Christian Sorg, Julia A. Camilleri, Angela R. Laird, Peter T. Fox, Sofie L. Valk, Simon B. Eickhoff, Masoud Tahmasian

## Abstract

**Background:** Neuroimaging studies have provided valuable insights into the macroscale impacts of antidepressants on brain functions in patients with major depressive disorder. However, the findings of individual studies are inconsistent. Here, we aimed to provide a quantitative synthesis of the literature to identify convergence of the reported findings at both regional and network levels and to examine their associations with neurotransmitter systems.

**Methods:** Through a comprehensive search in PubMed and Scopus databases, we reviewed 5,258 abstracts and identified 36 eligible functional neuroimaging studies on antidepressant effects in major depressive disorder. Activation likelihood estimation was used to investigate regional convergence of the reported foci of consistent antidepressant effects, followed by functional decoding and connectivity mapping of the convergent clusters. Additionally, utilizing group-averaged data from the Human Connectome Project, we assessed convergent resting-state functional connectivity patterns of the reported foci. Next, we compared the convergent circuit with the circuits targeted by transcranial magnetic stimulation (TMS) therapy. Last, we studied the association of regional and network-level convergence maps with selected neurotransmitter receptors/transporters maps.

**Results:** No regional convergence was found across foci of treatment-associated alterations in functional imaging. Subgroup analysis across the Treated > Untreated contrast revealed a convergent cluster in the left dorsolateral prefrontal cortex, which was associated with working memory and attention behavioral domains. Moreover, we found network-level convergence of the treatment-associated alterations in a circuit more prominent in the frontoparietal areas. This circuit was co-aligned with circuits targeted by “anti-subgenual” and “Beam F3” TMS therapy. We observed no significant correlations between our meta-analytic findings with the maps of neurotransmitter receptors/transporters.

**Conclusion:** Our findings highlight the importance of the frontoparietal network and the left dorsolateral prefrontal cortex in the therapeutic effects of antidepressants, which may relate to their role in improving executive functions and emotional processing.

## Introduction

Major depressive disorder (MDD) is the most common psychiatric disorder and a leading cause of disability worldwide [1]. Despite decades of research and the development of various pharmacological, psychological, and stimulation-based therapy, optimal treatment of MDD remains a challenge [2]. The conventional antidepressant medications, which are the mainstay of MDD treatment, can only achieve clinical response after several weeks of treatment [3] and only in around half the patients [4]. The challenges in the treatment of MDD are partly due to our limited understanding of the mechanisms by which antidepressants interact with the complex and heterogeneous neurobiology of MDD.

The monoamine neurotransmitter hypothesis of MDD postulates that decreased levels of serotonin and norepinephrine in specific brain regions are responsible for depressive symptoms, and antidepressant medications can normalize the imbalance in neurotransmitter levels [5]. While this hypothesis has dominated the field of MDD research and treatment for decades, it is increasingly being questioned, as the supporting evidence for a decreased concentration/activity of serotonin in MDD has been found inconclusive [6]. This, together with the discovery of rapid antidepressant effects of ketamine, a glutamate receptor antagonist [7, 8], suggests that the therapeutic effects of antidepressants cannot be simply explained as rebalancing the synaptic levels of the monoamine neurotransmitters. Thus, it is crucial to study the macroscale effects of antidepressant medications on the brain regions and networks beyond their neurochemical and cellular effects. Understanding these macroscale effects may help better understand their clinical effects on various symptoms of MDD, which is ultimately needed to improve treatment outcomes.

Neuroimaging techniques such as functional magnetic resonance imaging (fMRI) and positron emission tomography (PET) have been used to study the macroscale effects of anti-depressants on brain activity, metabolism, or connectivity [9, 10]. These studies have reported diverse functional effects of antidepressants on various brain areas, such as: i) normalizing reactivity of the amygdala, insula, and anterior cingulate cortex to negative stimuli [11–14], ii) reducing metabolism of paralimbic and subcortical areas, paralleled by increasing metabolism of fronto-parietal areas [15], or iii) modulation of resting-state function in the frontal, limbic and occipital areas as well as basal ganglia [16–18]. However, while individual neuroimaging studies are useful, the breadth of the literature and inconsistencies among the reported findings [10], necessitates a quantitative synthesis of the published literature to identify the most consistent findings that are robust to the large variability of the individual studies (in terms of clinical features and methodology), as well as their susceptibility to false positive/negative effects due to the usually small samples [19, 20].

Neuroimaging meta-analysis is a promising tool that enables a quantitative synthesis of the previously published literature [21, 22]. The most common approach in neuroimaging meta-analyses, i.e., coordinate-based meta-analysis (CBMA), aims to find potential regional convergence across the peak coordinates of the reported effects in individual studies [23]. Several neuroimaging meta-analyses have previously used this approach to study the regional convergence of the brain effects associated with the treatment of MDD, focusing on various therapeutic approaches and different neuroimaging modalities [24–30]. However, MDD is increasingly being recognized as a brain network disorder with distributed abnormalities across the whole brain, and similarly, the antidepressants’ effects could be distributed across the brain rather than localized [9]. Such distributed effects may be overlooked by the CBMA approaches, which are inherently intended for regional localization of effects. Recently, a novel meta-analytic approach has been introduced which aims to identify the convergence of reported findings at the level of networks by characterizing the normative convergent connectivity of the reported foci tested against random foci [31]. Using this approach, it was shown that despite a lack of regional convergence of reported abnormalities in MDD [32], there is a convergence of their connectivity in circuits which recapitulates clinically meaningful models of MDD [31].

Here, we aimed to identify how the findings of the previous functional neuroimaging studies on the effects of antidepressants converge on both regional and network levels by performing an updated CBMA as well as a network-level meta-analysis on the reported findings. Following, we compared our meta-analytic findings with the targets of transcranial magnetic stimulation (TMS) therapy and their associated circuits. Last, we asked whether the pattern of the observed meta-analytic effects of antidepressant medications on functional imaging can be potentially explained by the regional distribution of the neurotransmitter receptors/transporters (NRT) linked to these medications, leveraging the publicly available PET maps of neurotransmitter receptors and transporters [33].

## Methods

This meta-analysis was performed according to the best-practice guidelines for neuroimaging meta-analyses [21, 22] and is reported adhering to the Preferred Reporting Items for Systematic Reviews and Meta-Analyses (PRISMA) statement [34]. The protocol for this study was pre-registered on the International Prospective Register of Systematic Reviews (PROSPERO, CRD42020213202).

### Search and study selection

We searched the PubMed and Scopus databases to identify peer-reviewed eligible neuroimaging studies investigating the effects of antidepressants on MDD. The search was performed in July 2022, using the keywords reported in Table S1. In addition, we searched the BrainMap annotated database of neuroimaging experiments using Sleuth by setting the diagnosis to MDD and pharmacology to the antidepressants [35–38]. Further, to avoid missing any additional relevant studies, we traced the references of relevant neuroimaging reviews/meta-analyses. Next, the duplicated records were removed, and the resulting 5258 unique records were assessed for eligibility by two raters (S.RJ. and S.SN.) independently. The eligibility of records was assessed first using their titles or abstracts and then, for the potentially relevant records, by examining their full texts. Other authors (A. E., A. S., M. T.) resolved any disagreements between the main raters.

As suggested previously [21, 22], original studies were included if: 1) they studied patients with MDD, excluding patients with other major psychiatric or neurological comorbidities and adolescent or late-life patients, 2) the patients were treated with antidepressants, 3) the antidepressants effects on the function of gray matter structures were investigated using eligible neuroimaging modalities, i.e., functional magnetic resonance imaging (including task-based [tb-fMRI], resting-state [rs-fMRI] and arterial spin labeling [ASL-fMRI]), fluorodeoxyglucose positron emission tomography (FDG-PET), or single-photon emission computed tomography (SPECT), 4) the results of pre- vs. post-treatment, treated vs. placebo/untreated, or group-by-time interaction contrasts were reported as peak coordinates of significant clusters in standard spaces (Montreal Neurological Institute [MNI] or Talairach) or were provided by the authors at our request, 5) the analysis was performed across the whole brain, was not limited to a region of interest (ROI) or hidden ROI conditions such as small volume correction (SVC), as these approaches are biased toward finding significance in the respective areas, hence violating the assumption of ALE method that all voxels of the brain have a unified chance of being reported [21, 22], and 6) at least six subjects were included in each group (Fig. 1).

**Fig. 1.**
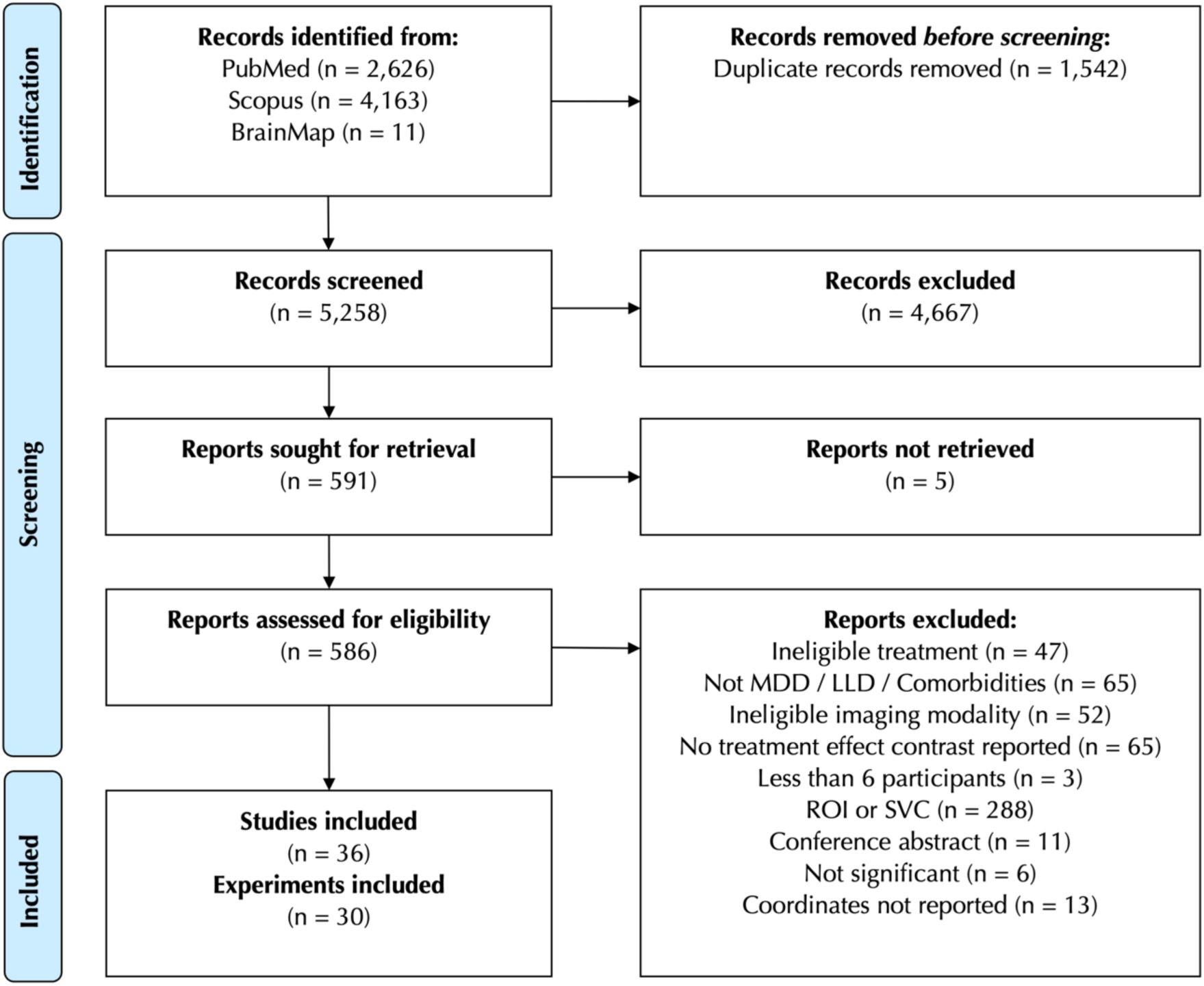
Study selection flowchart. MDD: major depressive disorder, LLD: late-life depression, ROI: region of interest, SVC: small volume correction.

### Data extraction and preprocessing

From the eligible studies, we extracted demographic and clinical data (number of participants, age, sex, response to treatment, medications, treatment duration), methodological details (imaging modality, scanner field strength, task paradigm, software package, statistical contrast, and the multiple comparisons correction method), as well as the peak coordinates/foci (x, y, z) of experiments’ findings. Of note, we use the term “study” to refer to an individual publication, and the term “experiment” to refer to the individual group-level contrasts reported within each “study” (e.g., Treated > Untreated). Following the data extraction, the coordinates reported in Talairach space were transformed into MNI space [39], so that all the experiments are in the same reference space. If the applied reference space was not explicitly reported or provided by authors after our request, we assumed the default settings of the software packages were used for normalization [21, 22]. In addition, to avoid spurious convergence over the experiments performed on the same/overlapping samples (reported within or across studies), in each meta-analysis, we merged the coordinates from multiple experiments pertaining to the same/overlapping samples, to make sure that each study contributes once per analysis, as suggested previously [21, 22, 40].

### Activation likelihood estimation

The revised version of the activation likelihood estimation (ALE) method [23] was used to test the regional convergence of the reported differences against the null hypothesis of randomly distributed findings across the brain. In this method, the peak coordinates were convolved with 3D Gaussian probability distributions that have a full width at half maximum, inversely proportional to the sample size. This allowed experiments with larger samples to have a greater statistical certainty in the meta-analysis. Next, for each experiment, the convolved foci were combined to generate per-experiment “modeled activation” (MA) maps. Subsequently, the MA maps for all the experiments included in the meta-analysis were combined into an ALE score map, representing the regional convergence of results at each location of the brain. The ALE score map was then statistically tested against a null distribution reflecting randomly distributed findings to distinguish true convergence from by-chance overlap [23, 40]. The resulting p-values were subsequently thresholded at voxel height threshold of p < 0.001. Finally, to avoid spurious findings [23], the resulting p-values were corrected for multiple comparisons using the family-wise error correction at the cluster level (cFWE). Specifically, the significance of cluster extents was assessed against a null distribution of maximum cluster sizes generated using a Monte Carlo approach with 10,000 permutations, wherein for each permutation ALE was performed on randomly selected foci distributed across the grey matter, and the maximum size of resulting null clusters were recorded.

In addition to an ALE meta-analysis on all included experiments (the ‘all-effects’ ALE) we performed several complementary ALE meta-analyses based on the direction of the effect (treatment contrast i.e., Treated > Untreated [Tr+] or Untreated > Treated [Tr-]), imaging modality, study design, and type of the antidepressants. The analyses were performed only if 12 or more experiments were included in each category, as ALE analyses with too few experiments are likely to be largely driven by a single experiment and, therefore, lack sufficient statistical power to provide valid results [41].

### Contribution assessment of the convergent clusters

For each significant convergent cluster, the relative contribution of included experiments was calculated as the fraction of the ALE values within the cluster accounted for by each experiment contributing to the cluster. Specifically, the contribution of each experiment to the cluster was calculated as follows: (i) Voxel-wise ratios were calculated between ALE score values of the cluster voxels after removing each experiment compared to the original ALE score calculated based on all included experiments. (ii) These ratios were averaged across the cluster voxels and subtracted from one, reflecting on average how much each experiment accounts for the ALE scores within the cluster. (iii) Contributions were normalized to a sum of 100%. Subsequently, we reported the contributions per categories of the contributing experiments according to modality, condition (task-based or resting-state), and medications, by summing up the contribution of the experiments within each category.

### Functional decoding of the convergent clusters

We applied the data from task-based functional neuroimaging experiments and their annotated behavioral domains (BD) included in the BrainMap database [35–38] to identify BDs significantly associated with the convergent clusters identified in the main ALE analyses [42]. In particular, we used binomial tests to assess whether the probability of each cluster activation given a particular BD, i.e., P(Activation|BD), is significantly higher than the overall a priori chance of its activation across all BDs, i.e., P(Activation). The resulting p-values were subsequently adjusted for multiple comparisons at false discovery rate (FDR) of 5%.

### Meta-analytic coactivation mapping of the convergent clusters

We investigated the task-based functional connectivity of the convergent clusters identified in the main ALE analyses using meta-analytic coactivation mapping (MACM) [43]. We used the data from task-based functional neuroimaging experiments on healthy individuals included in the BrainMap database [35–38]. For each identified convergence cluster from the main ALE analyses, we identified all the experiments that reported at least one focus of activation therein, and after merging the experiments reported within each study, performed an ALE meta-analysis across those studies, thresholded at p_cFWE_ < 0.05. This approach identifies brain regions consistently co-activated with the convergent cluster across all task-based functional neuroimaging experiments.

### Resting-state functional connectivity of the convergent clusters

We obtained the group-averaged dense resting-state functional connectivity (RSFC) matrix of the Human Connectome Project – Young Adults (HCP-YA) dataset (n=1003) available in *Cifti* format [44, 45]. The convergent cluster in the MNI space was transformed to *fsLR* space using *neuromaps* [46] and in turn mapped to *Cifti* ‘grayordinates’ (cortical vertex or subcortical voxel) covering the cluster extent. Subsequently, the whole-brain RSFC maps of these grayordinates were extracted from the HCP dense RSFC and were uniformly averaged, resulting in a RSFC map of the convergent cluster.

### Network-based meta-analysis

In addition to the conventional CBMA, we performed a network-based meta-analysis approach [31], to identify convergent functional connectivity of the reported foci compared to randomly distributed foci. We used the normative group-averaged dense RSFC matrix of the HCP dataset in these analyses. For the given set of experiments in the all-effects, Tr+ and Tr- analyses, we performed the following: (i) The MNI coordinates of all the reported foci in the included experiments were mapped from the voxel space to their closest grayordinate based on Euclidean distance. The foci with no grayordinate in their 10 mm radius were excluded (16 out of 528). Of note, the median distance of the mapped grayordinates from the MNI coordinates of foci was 2.26 mm. (ii) The whole-brain RSFC maps of the foci were extracted from the HCP dense RSFC and averaged in two levels: First, the RSFC maps of the foci from each experiment were averaged across the foci to create average experiment-specific RSFC maps. Second, average experiment-specific RSFC maps were averaged across experiments, weighted by their sample sizes, which resulted in a pooled RSFC map of all the experiments included in the analysis. (iii) This observed RSFC map was compared to a permutation-based null distribution of RSFCs to create a Z-scored convergent connectivity map. Specifically, in each of the 1000 permutations, we randomly sampled an equal number of foci as reported in the included experiments and averaged their RSFC maps as described above in step ii, resulting in a set of 1000 null RSFC maps. (iv) We calculated two-tailed p-values as the frequency of null pooled RSFC exceeding the observed pooled RSFC at each grayordinate, and subsequently transformed the p-values to Z scores. These maps, referred to as ‘convergent connectivity maps’, reflect greater- or lower-than-chance connectivity of the reported foci to the rest of the brain, which indicate convergent circuits connected to the locations of reported antidepressant treatment effects. Of note, to assess the effect of weighting the second-level average by experiment sample sizes, we additionally performed a sensitivity analysis in which this weighting was not applied.

Using a similar approach, we additionally assessed convergent connectivity of the reported foci to the seven canonical resting-state networks [47]. Here, we averaged the true and null pooled RSFC maps (calculated in steps ii and iii above) within each network. Following, for each of the seven networks, a two-tailed p-value was calculated as the frequency of null within-network average of pooled RSFC exceeding that of the true foci. Lastly, we corrected for multiple comparisons across the seven networks by FDR adjustment at 5%.

### Association with transcranial magnetic stimulation targets

We compared the location of the ALE convergent clusters with four TMS target coordinates, including the anatomical “5-CM” site (MNI -41, 16, 54) [48], “Beam F3” site (MNI -41, 42, 34) [49] and the “anti-subgenual” site (MNI –38, 44, 26) [50] used in clinical trials. In addition, we extracted the RSFC maps of grayordinates corresponding to these coordinates from the HCP dense RSFC and evaluated their spatial correlations with the all-effects convergent connectivity map of antidepressant effects as well as the RSFC maps of ALE convergent clusters.

In these correlations we parcellated the maps using Schaefer-400 parcellation in the cortex (400 parcels) and Tian S2 parcellation in the subcortex (32 parcels), and accounted for spatial autocorrelation by using permutation testing of variogram-based surrogated maps. In this approach, random surrogate maps were created with variograms that were approximately matched to that of the original map, as implemented in *BrainSMASH* [51]. Following, the Pearson correlation coefficient between observed maps *X* and *Y* was compared against a non-parametric null distribution of coefficients resulting from correlating surrogates of *X* with the observed map *Y*. Lastly, we corrected for multiple comparisons across the correlations using FDR adjustment at 5%. Notably, these associations were performed between parcellated maps given high computational costs of generating variogram-based surrogate maps at the level of grayordinates.

### Association of meta-analytic findings with neurotransmitter receptor/transporter densities

The PET maps of tracers associated with NRT were obtained from a previous study [33], which curated these maps from various sources [52–69]. These maps were based on tracers for serotoninergic and noradrenergic receptors/transporters (5HT1a, 5HT1b, 5HT2a, 5HT4, 5HT6, 5HTT, NAT) as well as the NMDA receptor. The PET maps were available in MNI volumetric space and were parcellated using Schaefer-400 parcellation in the cortex (400 parcels) and Tian S2 parcellation in the subcortex (32 parcels), and were subsequently Z-scored across parcels. In case multiple maps were available for a NRT we calculated an averaged map weighted by the sample size of the source studies.

We then calculated the correlation of parcellated NRT maps with the all-effects convergent connectivity map while accounting for spatial autocorrelation by using variogram-based permutation testing as described above. In addition, we tested for over-/under-expression of the NRTs in the ALE convergent clusters. To do so, we first normalized the Z-scored and parcellated maps of NRTs to 0-1 and after projecting them to the cortical surface, calculated the median normalized density of each NRT within the convergent cluster. Next, we compared the observed median densities against a null distribution calculated based on surrogate NRT maps with preserved spatial autocorrelation which were generated using *BrainSmash* [51]. In all tests, the resulting p-values were corrected for multiple comparisons across the NRT maps by using FDR adjustment at 5%.

### Data and code availability

The code used in this study can be accessed in a GitHub repository at https://github.com/amnsbr/antidepressants_meta. This repository includes all the Python code used to run the analyses and generate the figures apart from the MATLAB code used for behavioral decoding, which is available upon request. In addition, the repository includes the coordinates and processed data reported in our study, with the exception of: i) HCP-YA dense connectome is accessible to registered users at https://db.humanconnectome.org/, and ii) BrainMap dataset which is available as a file upon request, but is additionally searchable using Sleuth (https://www.brainmap.org/sleuth/).

## Results

### Experiments included in the meta-analysis

The study selection process is depicted in Fig. 1. We screened 5258 records resulting from our broad and sensitive search and assessed 586 full texts for eligibility to finally include 36 studies and 30 experiments with non-overlapping samples (Table 1 and Table S2) [12–18, 70–98]. Collectively 848 MDD patients were included in the experiments. The patients were treated using SSRIs (n=17), ketamine (n=7), SNRIs (n=7), mirtazapine (n=2), clomipramine (n=1), quetiapine (n=1), or bupropion (n=1). In six experiments, the patients received variable medications. The imaging modalities included were tb-fMRI (n=18), FDG-PET (n=4), H2O-PET (n=1), rs-fMRI (n=4), ASL-fMRI (n=2) and ^99m^Tc-HMPAO SPECT (n=2).

**Table 1.**
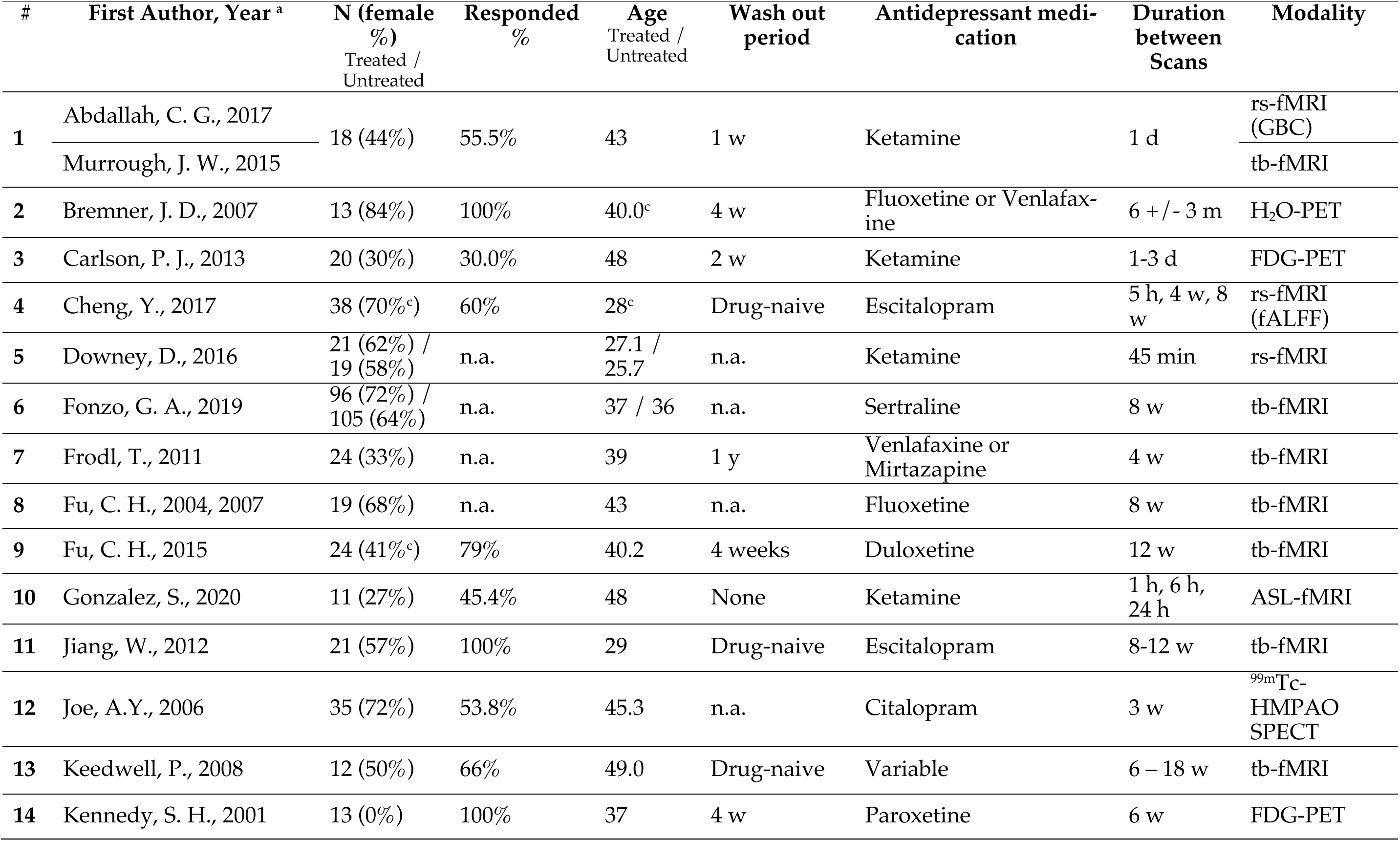
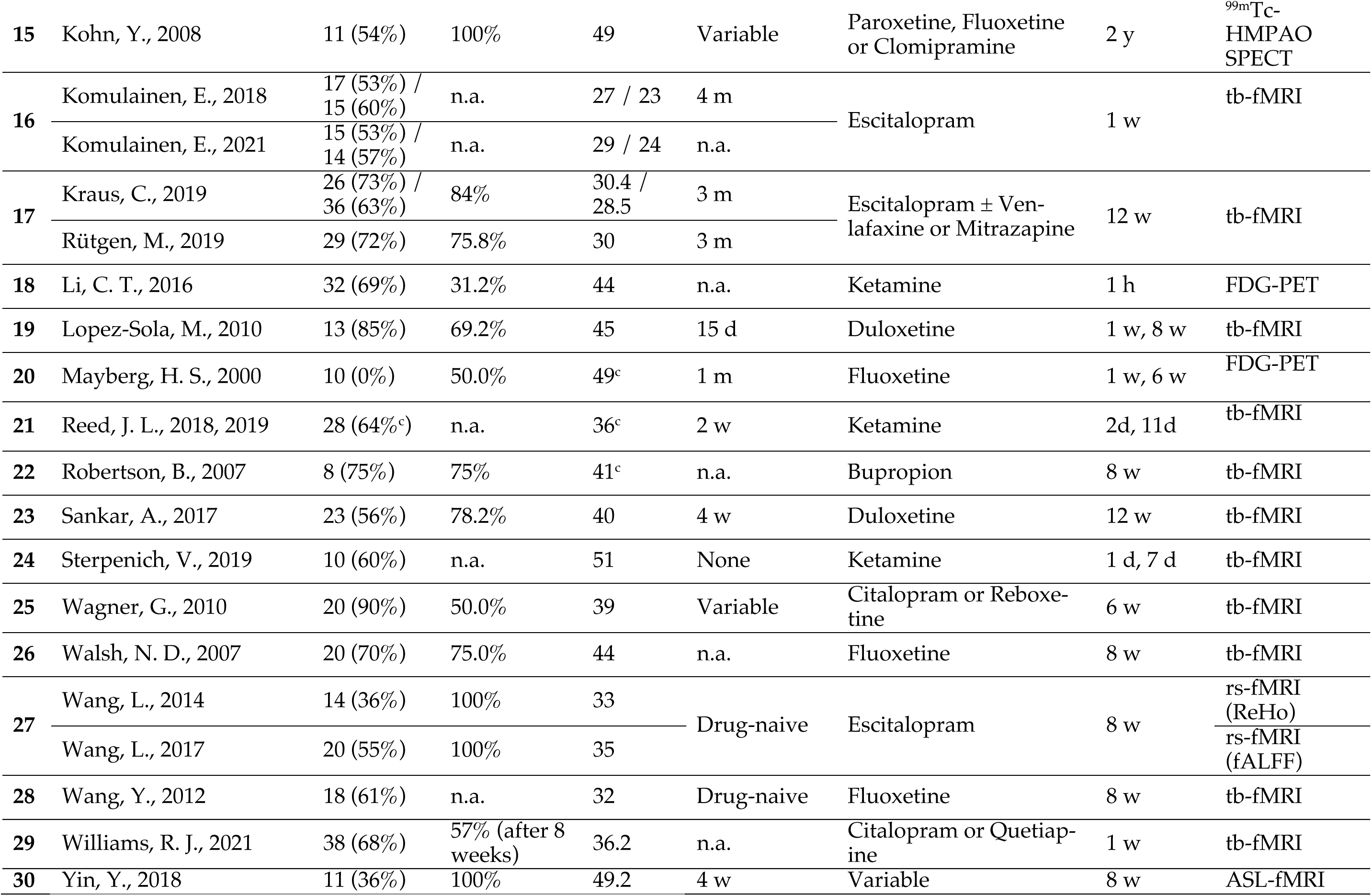
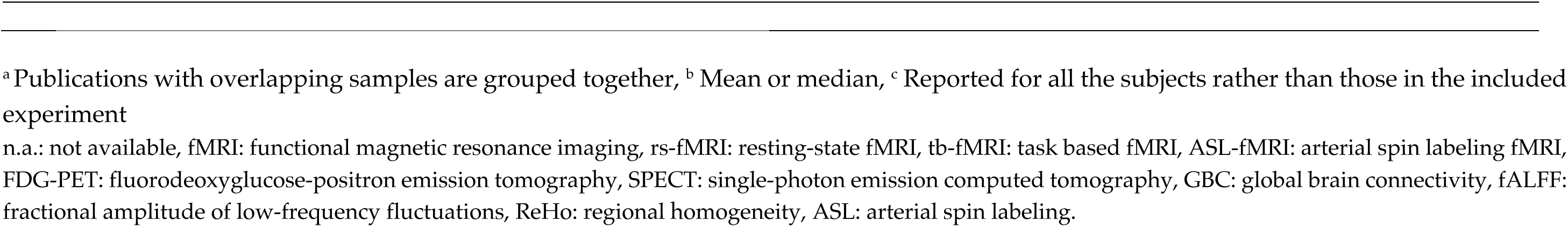
Characteristics of studies included in this meta-analysis.

### Convergent regional effects of antidepressants

No statistically significant regional convergence was found in our ALE meta-analysis on all included experiments reflecting treatment-related alterations in brain function (p_cFWE_ > 0.387), nor in its subgroup analyses limited to specific types of treatments or modalities (Table 2). However, among the Tr+ experiments (n=20) we observed a significant cluster of convergence in the left middle frontal gyrus within the dorsolateral prefrontal cortex (DLPFC) (MNI -38, 30, 28; 132 voxels) (Fig. 2). The convergence in this cluster was driven by contributions from seven experiments from eight studies [15, 17, 81–84, 86, 97]. The relative contribution of experiments using different medications included SSRIs (58.3%), ketamine (25.3%), and variable classes (16.4%). The contribution of experiments using PET (56.3%) was the highest, followed by fMRI (43.4%) and SPECT (0.3%). The scanning paradigms of contributing experiments included resting-state (81.0%) and emotional tasks (18.9%). Following, we investigated regional convergence of the Tr+ experiments across more specific subgroups of these experiments and found additional/different clusters (Fig. S1). To assess the convergence across classic antidepressants, we performed a subgroup analysis after excluding ketamine among 14 Tr+ experiments and found clusters of convergence in the left (MNI -38, 30, 30; 95 voxels) and right DLPFC (MNI 44, 26, 24; 106 voxels). In addition, to assess the effect of study design, a subgroup analysis was performed on 18 Tr+ experiments that only reported pre- versus post-treatment effects and revealed convergent clusters in the left (MNI -38, 30, 28; 135 voxels) and right DLPFC (MNI 44, 26, 24; 99 voxels). Following, we investigated the effect of treatment duration and performed an ALE on Tr+ effects reported after more than four weeks of treatment (12 experiments) which revealed a convergent cluster in the medial superior frontal gyrus (MNI 8, 54, 30; 104 voxels). Lastly, to assess the effect of variability in clinical outcomes, an ALE was performed on 12 Tr+ experiments which reported ≥ 50% rate of clinical response, and revealed a cluster of convergence in the left supramarginal gyrus (MNI -48, -44, 40; 123 voxels) in addition to the right DLPFC (MNI 44, 26, 24; 123 voxels).

**Fig. 2.**
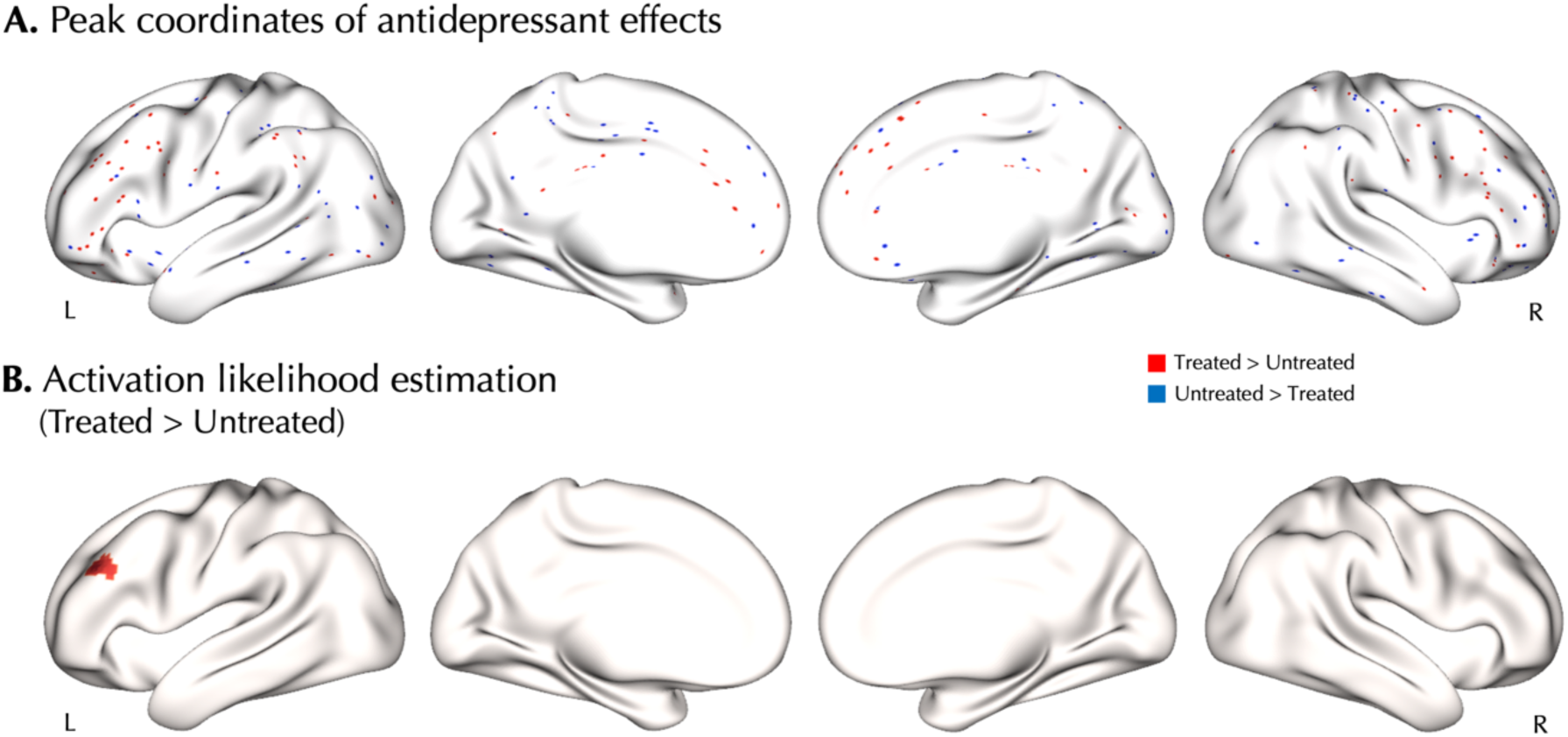
Treatment-induced increase of voxel-based physiology in the left dorsolateral prefrontal cortex. A. Peak coordinates of the included experiments in Treated > Untreated (red) and Untreated > Treated (blue) comparisons. Each dot represents a peak coordinate. B. Activation likelihood estimation showed significant convergence of Treated > Untreated comparisons in the left dorsolateral prefrontal cortex (DLPFC) after family-wise error correction at cluster level.

**Table 2.** Activation Likelihood Estimation (ALE) analyses on the effects of antidepressants in major depressive disorder.

| Experiments | Comparison | N | Min $p_{cFWE}^a$ | Convergence |
| --- | --- | --- | --- | --- |
| All | All | 30 | 0.387 | - |
|  | Treated > Untreated | 20 | **0.005** | L DLPFC |
|  | Treated < Untreated | 21 | 0.615 | - |
| *Based on modality* |  |  |  |  |
| Rest | All | 12 | 0.052 | - |
| Task | All | 19 | 0.970 | - |
| *Based on treatment and clinical setting* |  |  |  |  |
| Excluding ketamine | All | 23 | 0.486 | - |
|  | Treated > Untreated | 14 | **0.024** | L DLPFC, R DLPFC |
|  | Treated < Untreated | 16 | 0.464 | - |
| Pre- versus post-treatment effects | All | 24 | 0.294 | - |
|  | Treated > Untreated | 18 | **0.004** | L DLPFC, R DLPFC |
|  | Treated < Untreated | 20 | 0.469 | - |
| Treatment duration > 4 weeks | All | 20 | 0.472 | - |
|  | Treated > Untreated | 12 | **0.010** | R mSFG |
|  | Treated < Untreated | 14 | 0.189 | - |
| Response in $\geq 50\%$ of subjects | All | 19 | 0.614 | - |
|  | Treated > Untreated | 12 | **0.007** | L SMG, R DLPFC |
|  | Treated < Untreated | 15 | 0.357 | - |

<sup>a</sup> Bold p-values indicate statistical significance.
cFWE: cluster-wise family-wise error, SSRI: selective serotonin reuptake inhibitor, L: left, R: right, DLPFC: dorsolateral prefrontal cortex, mSFG: medial superior frontal gyrus, SMG: supramarginal gyrus.

We observed no significant convergence in the ALE meta-analysis performed on the opposite contrast among Tr- experiments (N = 21; p_cFWE_ > 0.615) as well as their more specific subgroups (Table 2).

### Functional decoding and MACM of the dorsolateral prefrontal cortex cluster

Next, we studied the behavioral relevance as well as task-based and resting-state FC of the convergent cluster identified in the main ALE meta-analysis on Tr+ experiments within the left DLPFC. Using the data from the BrainMap database, we observed that the behavioral domains of working memory (likelihood ratio = 1.85) and attention (likelihood ratio = 1.43) were significantly associated with this cluster’s activation, yet they did not survive FDR correction.

The MACM of the left DLPFC cluster showed its significant co-activation with regions in the prefrontal cortex, superior parietal lobule, insula, and anterior cingulate and paracingulate cortices (p_cFWE_ < 0.05; Fig. 3A). The average RSFC of the left DLPFC cluster based on the HCP dataset dense connectome showed its connectivity with widespread regions in the pre-frontal cortex, superior frontal gyrus, insula, anterior cingulate, paracingulate cortices, supramarginal gyrus, inferior temporal gyrus, and basal ganglia, and its anti-correlation with regions in the subgenual anterior cingulate cortex, orbitofrontal cortex, posterior cingulate, angular gyrus, and temporal pole (Fig. 3B).

**Fig. 3.**
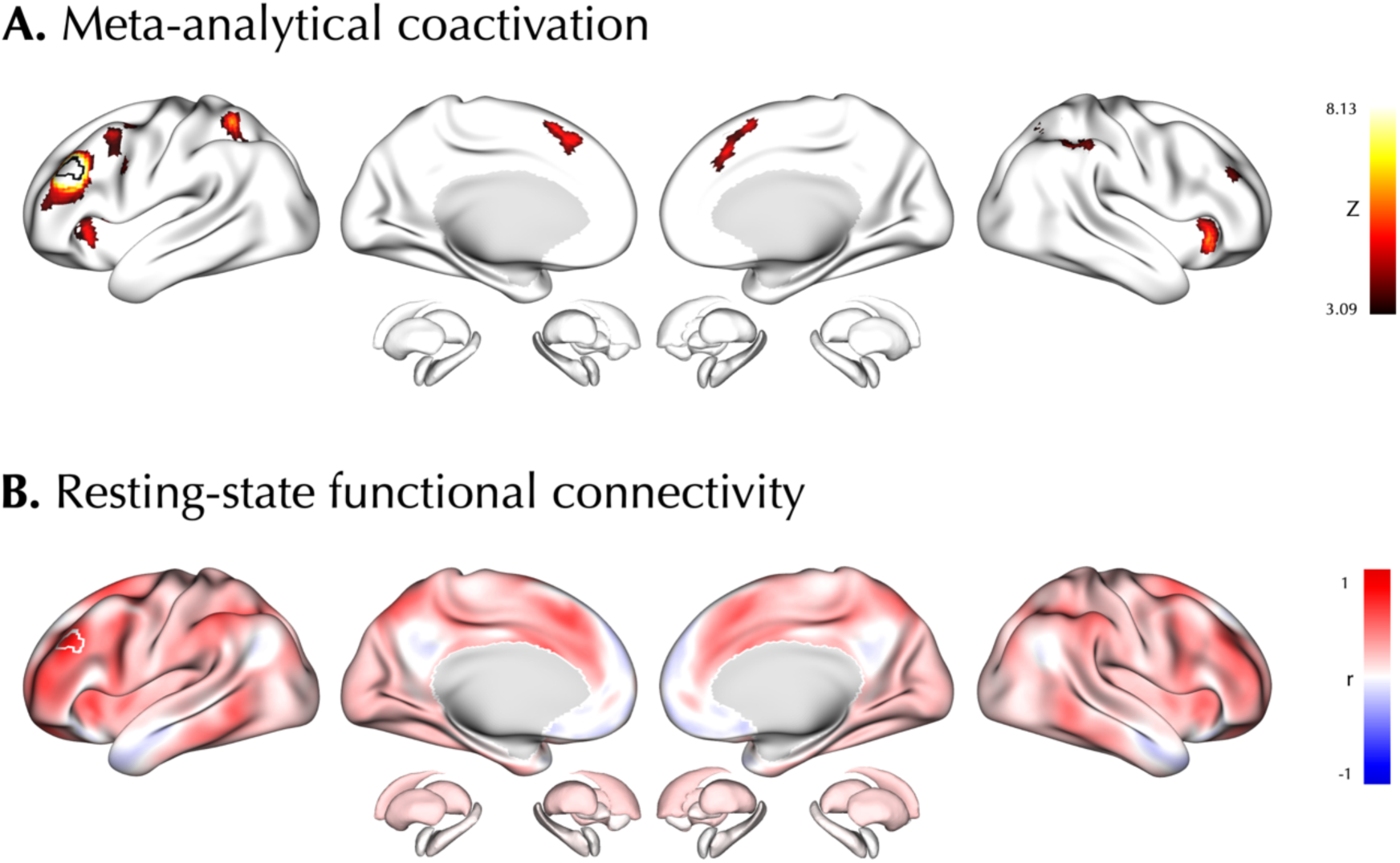
Connectivity mapping of the left dorsolateral prefrontal cortex cluster. Using the center of convergent cluster at the left dorsolateral prefrontal cortex as the seed (outlined patch), the meta-analytical co-activation (A) and resting-state functional connectivity (B) maps are shown.

### Convergent connectivity mapping of antidepressants’ effects

MDD and its treatment are believed to affect distributed regions and networks in the brain [9], and ALE, which aims to identify *regional* convergence of localized effects cannot characterize convergence of such *distributed* effects. Therefore, we next aimed to investigate the meta-analytic effects of antidepressants at a network level, following a recently introduced approach [31]. To do so, we used the group-averaged dense functional connectome obtained from the HCP-YA dataset and quantified the convergent connectivity of the reported coordinates of antidepressant effects, which was compared against null connectivity patterns of random points.

The peak coordinates of all the included experiments, indicating alterations in functional imaging measures associated with antidepressants (512 foci from 30 experiments), showed greater-than-chance connectivity of these coordinates with widespread regions in the dorsolateral and medial prefrontal cortex, anterior insula, precuneus, supramarginal gyrus and inferior temporal gyrus. At the level of canonical resting-state networks, these foci showed significant greater-than-chance convergent connectivity to the frontoparietal (FPN; <Z> = 0.23, p_FDR_ < 0.05) network (Fig. 4). The convergent connectivity described above was calculated by taking an average of pooled RSFC maps across experiments which was weighted by their sample sizes. Subsequently, in a sensitivity analysis we showed a similar convergent connectivity map when this weighting was not applied (r = 0.922, p_variogram_ < 0.001; Fig. S2), though at the level of canonical resting-state networks it revealed no significant effects.

**Fig. 4.**
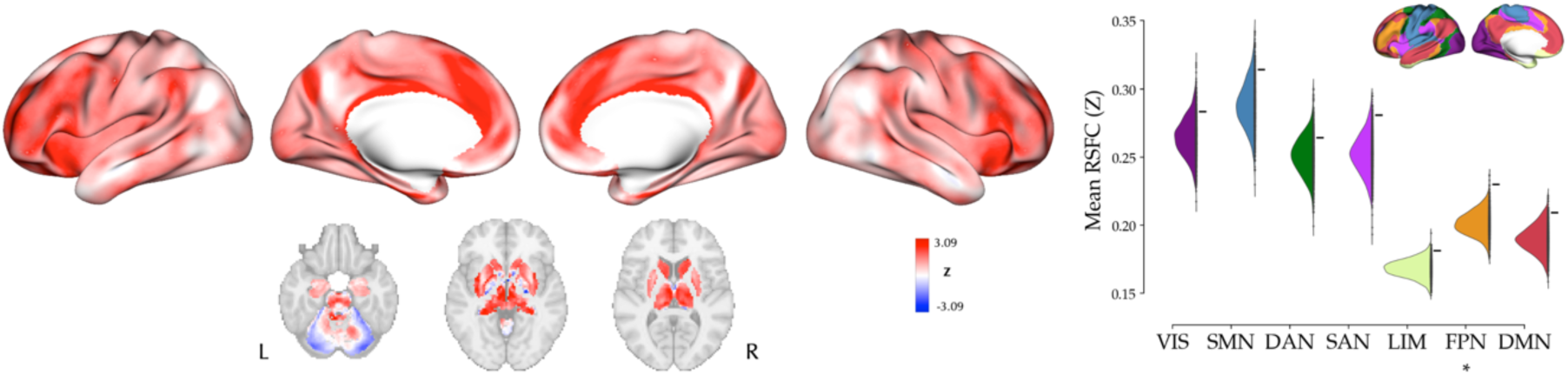
Convergent connectivity mapping of antidepressant effects. *left*: The cortical and subcortical map represent Z-scored convergent connectivity map of the foci from all experiments. *right:* Mean resting-state functional connectivity (RSFC) of the observed foci across canonical resting-state networks (denoted by “-“) compared against null mean values calculated based on 1000 per-mutations of randomly selected foci (half-violin plots). In frontoparietal network (FPN) observed mean RSFC was significantly more extreme than the null distribution in a two-tailed test. VIS: visual network, SMN: somatomotor network, DAN: dorsal attention network, SAN: salience network, LIM: limbic network, FPN: frontoparietal network, DMN: default mode network.

The network-level analyses separately performed on the Tr+ (177 foci from 20 experiments) and Tr- effects (206 foci from 21 experiments) revealed convergent connectivity maps which were anti-correlated with each other (r = -0.57, p_variogram_ < 0.001; Fig. S3). The convergent connectivity map of all experiments (Fig. 4) was significantly correlated with the convergent connectivity map of Tr+ (r = 0.48, p_variogram_ < 0.001) but not Tr- experiments (r = 0.051, p_variogram_ = 0.732). Furthermore, the contrast-based analyses at the level of canonical resting-state networks revealed no significant greater-/lower-than-chance connectivity of the foci with these networks after FDR correction.

### The spatial localization of antidepressants’ effects compared with TMS targets

The left DLPFC is one of the most common stimulation targets in the TMS treatment of MDD [99, 100]. Next, we explored whether our meta-analytic findings on the convergent effects of antidepressants might spatially correspond with the different TMS targets. We compared the location of the peak coordinate of the left DLPFC cluster identified in the Tr+ ALE meta-analysis and observed its distance with the different targets ranged from 13 to 27 mm (Fig. 5A). Moreover, the RSFC map of the Tr+ cluster showed significant correlations (p_variogram, FDR_ < 0.001), positively with the RSFC maps of “Beam F3” (r = 0.82) and “anti-subgenual” (r = 0.72) targets, but negatively with the RSFC map of “5-CM” (r = -0.50) target (Fig. 5B). In addition, the convergent connectivity map of antidepressant effects showed a significant and positive correlation with the RSFC of “anti-subgenual” target (r = 0.31, p_variogram,FDR_ < 0.001).

**Fig 5.**
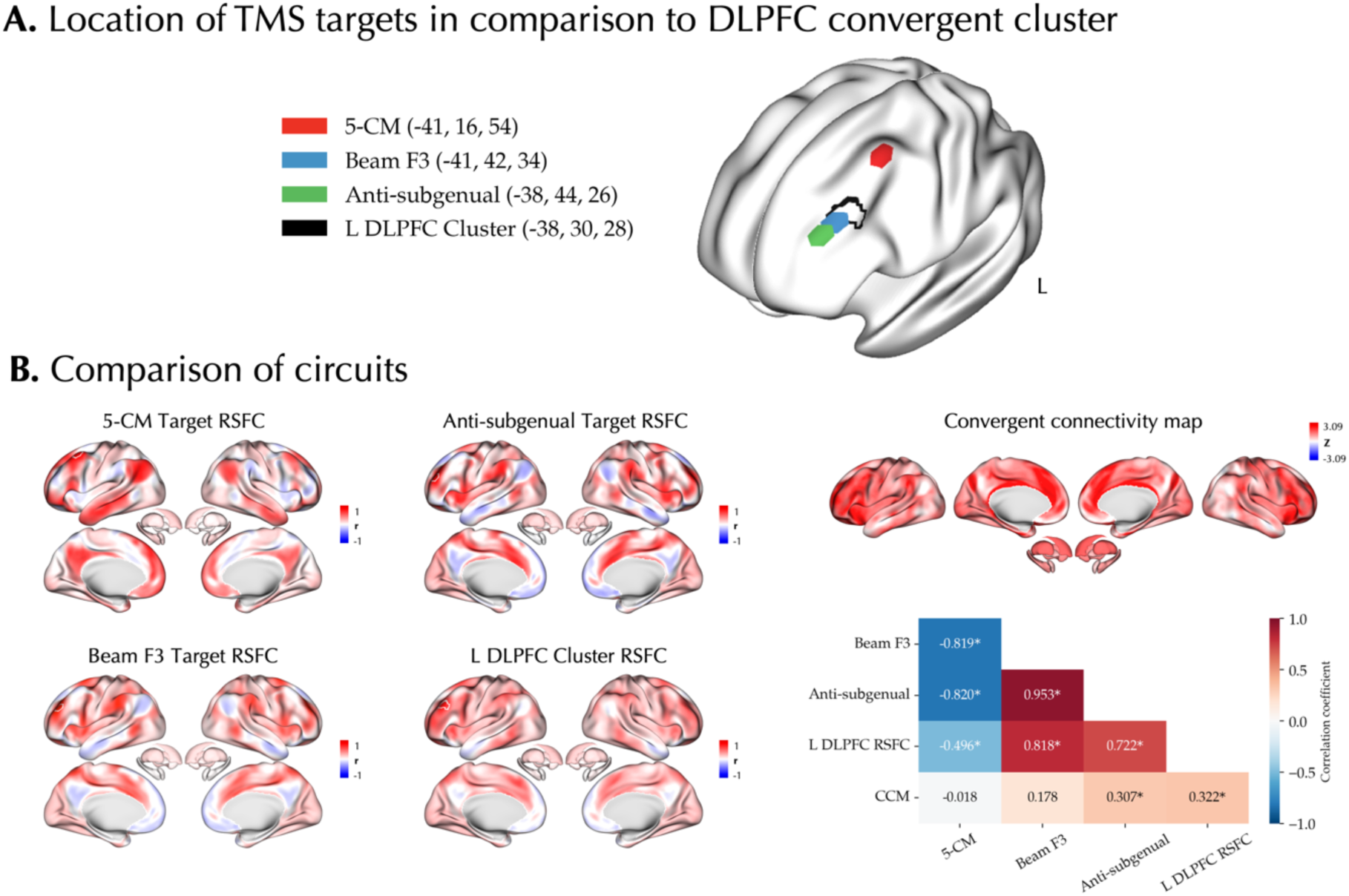
Association of antidepressants meta-analytic effects and transcranial magnetic stimulation targets. **A.** The locations of three different TMS targets is shown in comparison to the left DLPFC convergent cluster identified in the activation likelihood estimation meta-analysis on Treated > Untreated experiments. **B.** The convergent connectivity map of antidepressant effects, the RSFC map of the left DLPFC cluster, and the RSFC maps of the different TMS sites as well as their cross-correlations are shown. In the correlation matrix within each cell the Pearson correlation coefficient is reported. Asterisks denote p_variogram, FDR_ < 0.05.

### The association between neurotransmitter receptor/transporter maps and meta-analytic effects of antidepressants

Lastly, we explored whether the regional and network-level convergence of antidepressant effects co-localizes with the spatial distribution of serotoninergic and noradrenergic NRTs as well as NMDA receptor (Fig. 6A) [33]. We first focused on the cluster of convergence of Tr+ effects in the left DLPFC and quantified the median density of each NRT (normalized to a range of [0–1]) in this region, showing the varying density of the NRTs. However, after FDR correction none of the NRTs were significantly over-/under-expressed in this cluster (Fig. 6B). Next, we evaluated the correlation of the parcellated convergent connectivity map with the NRT maps and observed no significant correlations after FDR correction and while accounting for the spatial autocorrelation (Fig. 6C).

**Fig. 6.**
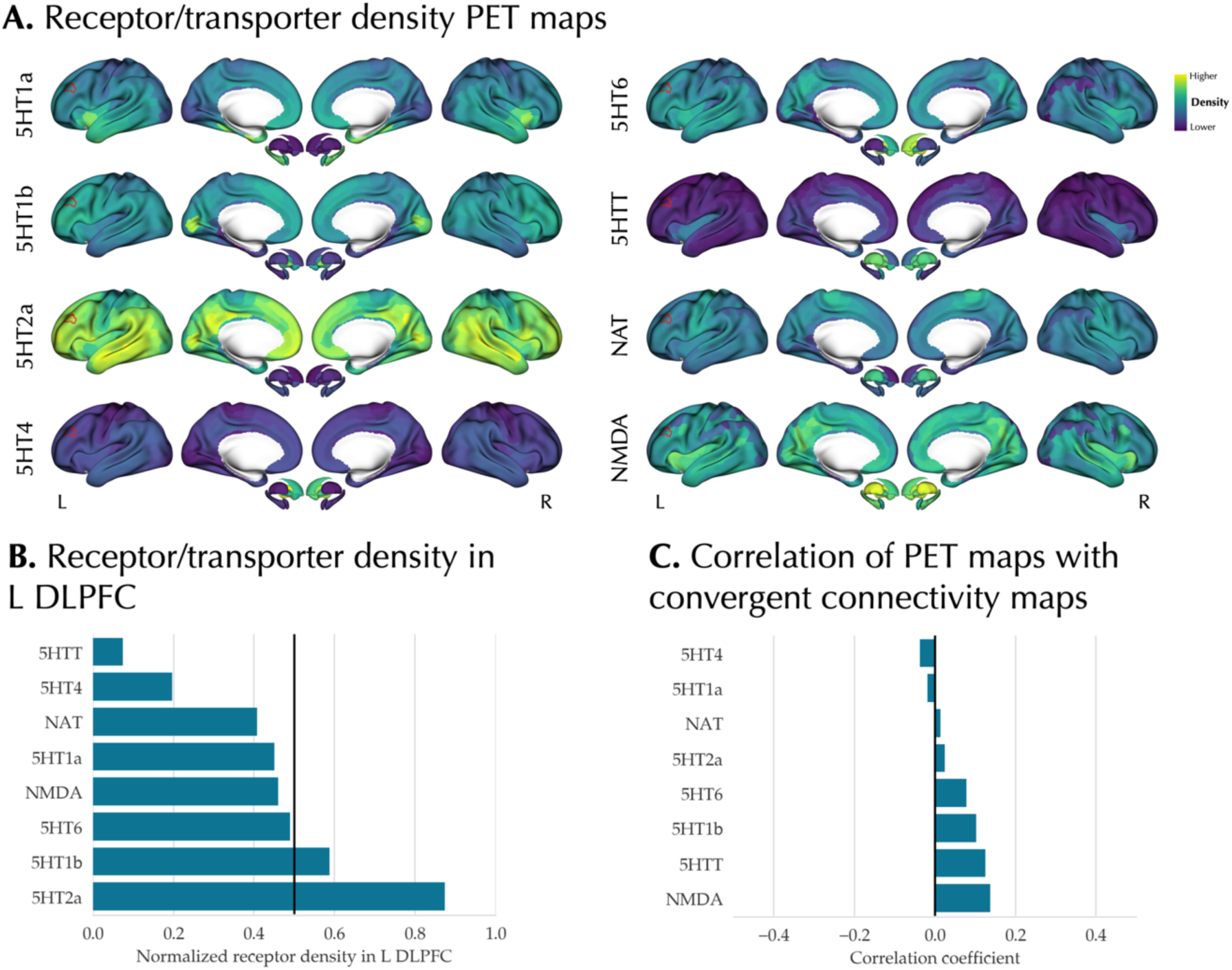
Association of meta-analytic findings with neurotransmitter receptor/transporter maps. **A.** The parcellated and Z-scored PET maps of neurotransmitter receptor/transporter (NRT) are shown. Red outline indicates the left dorsolateral prefrontal cortex (L DLPFC) convergent cluster identified in the activation likelihood estimation meta-analysis on Treated > Untreated experiments. **B.** Median normalized density of NRTs in L DLPFC cluster. None of the NRTs showed significantly more extreme normalized density in this cluster compared to a null distribution created using variogram-based surrogate maps and after false discovery rate adjustment. **C.** Pearson correlation of parcellated convergent connectivity map with the NRT maps. None of the correlations were significant using variogram-based surrogates and after false discovery rate adjustment.

## Discussion

In the present study, we synthesized findings of the neuroimaging literature on the brain effects associated with pharmacotherapy of MDD at regional and network levels. At the regional level, our meta-analysis showed no significant convergence across all the included experiments, though we found convergence of the reported treatment-associated increases of functional measures in the left DLPFC. This convergent cluster was associated with working memory and attention behavioral domains and showed meta-analytical connectivity with regions in the prefrontal cortex, superior parietal lobule, and insula. Extending our meta-analysis to the network level, we found greater-than-chance convergent connectivity of the reported foci of antidepressant effects primarily to the frontoparietal network. Subsequently, we found that the convergent connectivity of the foci was co-aligned with circuits connected to the “anti-subgenual” and “Beam F3” TMS targets. Last, we did not observe any significant association between the spatial pattern of the regional and network-level meta-analytic effects and the NRT maps.

### Beyond regional convergence: Network-level convergent effects of anti-depressants to the frontoparietal network

The pathology in MDD is increasingly thought to be distributed across brain regions and networks, rather than solely being localized [9]. Previous ALE meta-analyses aimed at identifying the *regional* convergence of abnormalities in MDD have revealed minimal or no regional convergence [32, 101, 102]. However, a recent meta-analysis by Cash *et al.* [31] revisited the functional imaging literature on brain abnormalities in MDD, aiming to identify the *network-level* convergence of abnormalities in MDD. They identified circuits of convergent connectivity related to the location of emotional and cognitive processing abnormalities in MDD, including lower-than-chance connectivity of the emotional processing abnormalities to DLPFC and pre-supplementary motor area, and greater-than-chance connectivity of the cognitive processing abnormalities to DLPFC, cingulum, insula, and precuneus [31]. Here, we found regional convergence of Tr+ functional effects in left DLPFC, yet our main ALE meta-analysis across all functional treatment effects revealed no significant regional convergence. The lack of regional convergence in our overall ALE meta-analysis, similar to the case of previous ALE meta-analyses on MDD disease effects, may be attributed to the biological and clinical heterogeneity of MDD and antidepressant effects, as well as the methodological heterogeneity of the included experiments [102]. But more importantly, it may reflect *distributed* rather than *localized* effects of antidepressants [9]. Therefore, following a similar approach to Cash et al. [31], we investigated *network-level* convergence of the reported antidepressant effects, and found their greater-than-chance connectivity in a network most prominent in the dorsolateral and medial prefrontal cortex, anterior insula, precuneus, supramarginal gyrus and inferior temporal gyrus. This circuit largely resembles the circuit of cognitive processing abnormalities in MDD [31]. Furthermore, at the level of canonical resting-state networks we found significant convergent connectivity of the reported findings with the FPN.

The convergent connectivity of reported antidepressant effects to the FPN, together with the cluster of regional convergence found in the ALE meta-analysis on the Tr+ experiments, highlights the importance of FPN and DLPFC in the therapeutic effects of antidepressants. These regions play pivotal roles in higher executive and cognitive functions of the brain, which are shown to be impaired in patients with MDD [103–106]. Indeed, more severe executive dysfunctions are linked with higher severity of depressive symptoms [105]. The executive and cognitive dysfunction in MDD is thought to contribute to emotional dysregulation, which is a hallmark of MDD psychopathology [107, 108]. Specifically, patients with MDD might have impairments in cognitive control when processing negative emotions, deficits in the inhibition of mood-incongruent material, and difficulties in attentional disengagement from negative stimuli, which are among the mechanisms that are thought to contribute to emotional dysregulation [107, 108]. Indeed, antidepressants medications are shown to improve the executive functioning of patients with MDD, in the domains of attention and processing speed [109], psychomotor speed [110] and cognitive interference inhibition [106], and can lead to better emotional regulation strategies [111]. Hypoactivity of the prefrontal cortex in MDD is thought to contribute to the deficits of executive functioning [105, 112, 113] and emotional regulation [108, 112, 114, 115]. For instance, patients with MDD have shown a reduced activity of the DLPFC during an attentional interference task using emotional distracters [116], which can be normalized by antidepressants [117]. Furthermore, the FPN in patients with MDD shows reduced within-network connectivity and decreased connectivity with the parietal regions of the dorsal attention network, as reported by a meta-analysis on seed-based RSFC studies [118]. In addition, hypoconnectivity of the FPN with the rest of the brain has been observed in relation to depressive symptoms in the general population [119]. The treatment of MDD using various therapeutic approaches can affect intra- and inter-network connectivity of the FPN [9], DMN [120], and SAN [121–123]. Overall, MDD is characterized by altered function and connectivity of distributed networks, importantly including the FPN, but also the SAN, DMN and limbic networks, which can be modulated by the treatment (see review in Chai et al. [9]).

### A similar network may be modulated by both antidepressants and TMS

The importance of DLPFC and FPN in MDD treatment has further been observed in nonpharmacological therapeutic approaches. For example, psychotherapy of patients with MDD and PTSD is shown to normalize the activity of DLPFC and increase the within-network connectivity of FPN [124]. In addition, the left DLPFC is one of the most common targets of stimulation in TMS therapy of MDD [99, 100]. High-frequency TMS applied to this region alters its activity, which in turn may have therapeutic effects by modulating the activity of its connected circuits [48, 99]. Interestingly, in our study, we found that both the convergent connectivity map of the antidepressant effects and the RSFC map of the left DLPFC cluster of convergence across Tr+ experiments were positively and significantly correlated with the “anti-subgenual” target RSFC map [50, 99, 100, 125]. Hyperactivity of the subgenual anterior cingulate cortex in MDD is thought to contribute to increased processing of negative stimuli [113]. Therefore, both the “anti-subgenual” TMS and antidepressant treatment of MDD might modulate the activity of a similar circuit, including DLPFC and sgACC. Of note, we found that the “5-CM” TMS target’s circuit was anticorrelated with the circuits of “anti-subgenual” and “Beam F3” TMS targets as well as our Tr+ convergence cluster in left DLPFC. Indeed, distinct circuits connected to these different TMS targets has been previously reported, and in a retrospective study, were found to relate to clinical response in distinct clusters of depressive symptoms [125].

### Lack of association between neurotranmitters and the system-level effects of antidepressants

The neurotransmitter hypothesis of MDD suggests that the dysregulation of the monoaminergic neurotransmitter systems is central to the pathophysiology of MDD, and antidepressants normalize the dysregulations of these neurotransmitter systems [5, 126, 127]. In our analyses, we found that the PET-based maps of serotoninergic and noradrenergic receptors and transporters were not significantly correlated with the regional and network-level meta-analytic effects of antidepressants. This suggests a disparity between the antidepressant effects on brain function, as observed in functional imaging studies, and the regions where their target NRTs are highly expressed. The observed divergence raises the question of what mechanisms may relate the micro-scale actions of antidepressants on the NRTs to their system-level effects on brain function. Molecular imaging techniques combined with functional imaging might provide clues to this link. The findings of molecular imaging studies in MDD and its treatment are diverse (see a comprehensive review by Ruhé et al. [127]). For example, there has been some evidence of decreased serotonin synthesis rate in the prefrontal and cingulate cortex of patients with MDD [128–130]. However, a recent umbrella review summarizing the research on the serotonin hypothesis of MDD concluded that there is a lack of convincing evidence for the association of MDD with serotoninergic deficits, such as a lower serotonin concentration or changes in the receptors [6]. Moreover, the antidepressive effects of ketamine, an NMDA receptor antagonist, highlight the importance of the other non-monoaminergic neurotransmitters in the pathophysiology and treatment of MDD [7, 8]. These findings suggest that although the monoaminergic neurotransmitter hypothesis of MDD helped us understand the pathophysiology of MDD, it may not provide a full understanding of the disease [6, 131, 132].

### An overview of neuroimaging meta-analyses on MDD treatments

The neuroimaging effects of treatment in MDD have been previously investigated in a number of CBMAs [24–30]. These studies have focused on various types of treatment, with more specific (e.g., only SSRI medications [27]) or broader (e.g., pharmacotherapy, psychotherapy, and electroconvulsive therapy [29]) scopes compared to our study. In addition, various imaging modalities under different conditions have been investigated, from focusing on fMRI experiments during emotional processing tasks [30] to a broader multimodal investigation of neuroimaging experiments [29]. Given the differences in the scope and methodology of the previous CBMAs, it is not surprising that they have reported variable findings (Table 3). However, it is important to note that according to the current guidelines [21, 22], there are a few methodological issues to consider in some of the (earlier) CBMAs, which may have influenced their findings. These issues include: i) a small number of experiments included in the main or subgroup analyses, which can limit the power and increase the risk of a single experiment dominating the findings [41], ii) including explicit or hidden ROI-based experiments which are biased to inflate significance in the selected brain area, iii) using less stringent methods of multiple comparisons correction, e.g., thresholding clusters simply by applying a lenient cluster extent and height, or by using FDR, or iv) performing ALE using the earlier versions of GingerALE (< 2.3.3), in which a software bug was reported that can lead to more lenient multiple comparisons correction [133]. Here, we set out to avoid such methodological issues by following the best-practice guidelines to conduct CBMAs [21, 22]. Furthermore, we provided network-level accounts of the effects of antidepressants reported in the literature [31], which acknowledges that the effects may be distributed rather than localized, and in doing so, complements the conventional CBMA approach of identifying regional convergence.

**Table 3.**
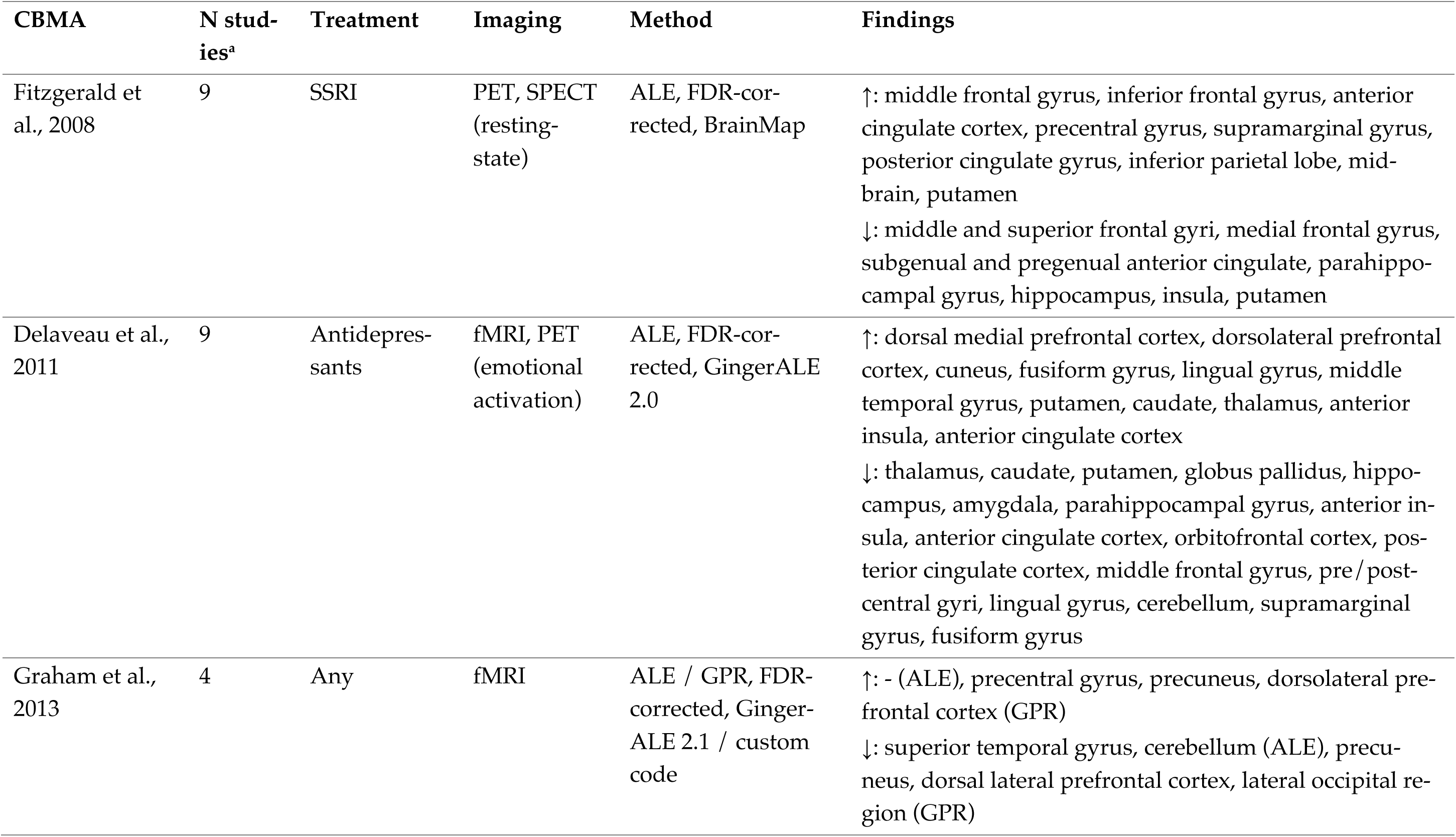
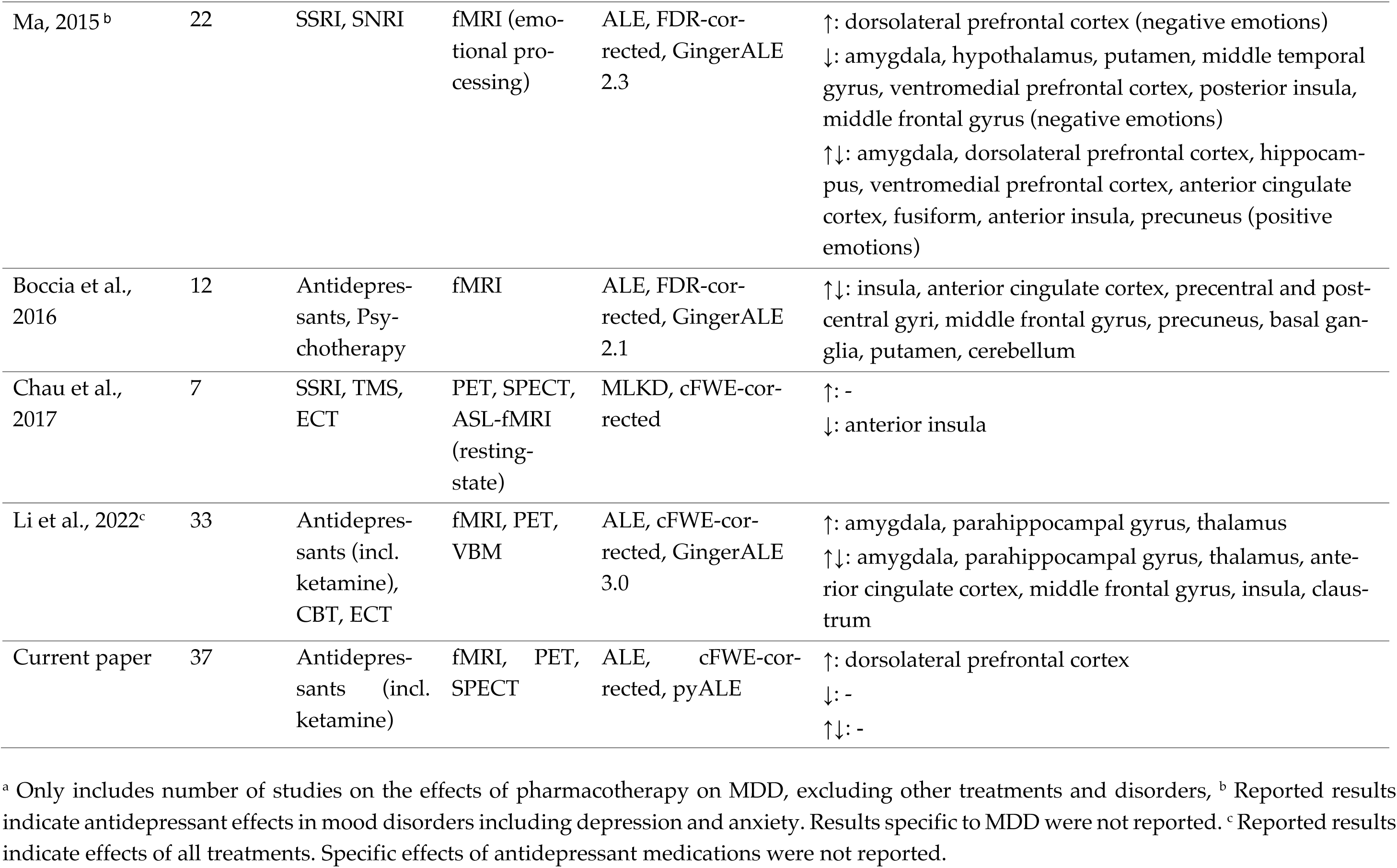
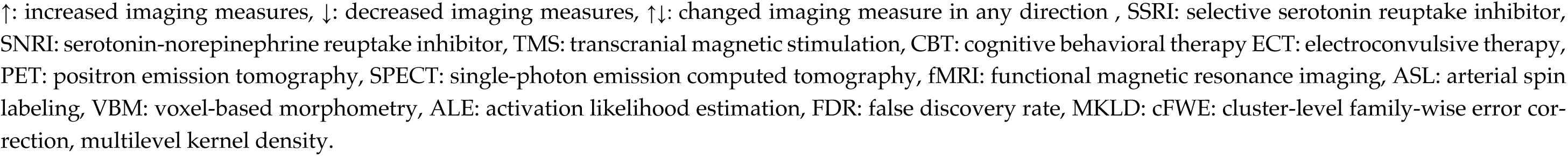
Comparison of the existing coordinate-based neuroimaging meta-analyses on the brain effects of antidepressants.

### Future directions and limitations

In this section, we highlight areas for future research that can extend our findings and address some of our limitations. First, while our study characterizes regional and network-level convergence of antidepressant effects across multiple imaging modalities, treatments, and clinical phenotypes, more focused meta-analyses will be needed on specific selections of experiments, when more data is available in the future. Relatedly, and on the other hand, broader meta-analyses on multiple treatment modalities (including medications, psychotherapy, and brain stimulation) can extend previous work [24, 29] and address the question of whether there is a general MDD treatment effect that is shared across different therapies. Second, we studied the neuroimaging effects of antidepressant medications on patients with MDD who had received but not necessarily responded to the treatment. There is considerable variability in the clinical outcomes of antidepressant treatment [4], and this is likely to be associated with variable neuroimaging effects. To investigate the heterogeneity of imaging effects related to the clinical outcomes at the level of included studies, we performed subgroup analyses focused on experiments reporting clinical response in at least half the patients. Among the Tr+ subset of those experiments, in a small ALE, we observed convergent clusters in the left supramarginal gyrus and the right DLPFC. However, further in-depth original and meta-analytic neuroimaging studies are needed to evaluate the inter-individual variability of treatment-induced changes in brain function and its relevance to clinical response, for example, by comparing the treatment-associated functional alterations between responders and non-responders. Additionally, utilizing multivariate machine learning models on imaging, genetics, and clinical data could provide valuable insights into the inter-individual variability among MDD patients in terms of disease characteristics and in turn, response to treatment, which is critical towards personalized treatment [134–136]. Third, as we did not restrict our data to randomized controlled trials, the included subjects may have received other interventions such as psychotherapy, which may confound the results. This issue can be addressed in a future meta-analysis focused on randomized controlled trials, when more studies are available. Fourth, our network-level meta-analysis was performed using a group-averaged functional connectome based on resting-state imaging data of healthy subjects, and therefore, provides an indirect view on network-level actions of antidepressants. Further large-scale studies are needed to investigate these effects using individual-level connectomic approaches on MDD patients treated with antidepressants.

## Conclusion

This comprehensive meta-analysis of the functional neuroimaging studies on the regional and network-level convergence of the effects of antidepressant medications in MDD underscores the importance of the FPN and particularly DLPFC. This observation may be attributed to the key roles of these regions in executive functions and emotional processing. The convergent regional and connectivity maps of antidepressant effects engaged circuits similar to those of “anti-subgenual” and “Beam F3” TMS targets, which may indicate common circuits are targeted by these different treatment modalities. Lastly, we identified no associations between our regional and network-level meta-analytic findings with the spatial maps of neurotransmitter receptors/transporters, suggesting that the localized functional effects of antidepressants cannot be directly explained by the localization of these receptors/transporters. Our study highlights the need for future research integrating multiple levels of antidepressant actions at the micro- and macroscale in the context of inter-individual variability of patients with MDD and their heterogeneous clinical outcomes.

## Data Availability

All data produced are available online. The coordinates of the foci reported in the included experiments are available at https://doi.org/10.6084/m9.figshare.24592539. The group-averaged dense resting-state functional connectivity matrix from the Human Connectome Project can be accessed at https://db.humanconnectome.org.

## Acknowledgements

AS and SLV were funded by the Max Planck Society (Otto Hahn award) and Helmholtz Association’s Initiative and Networking Fund under the Helmholtz International Lab grant agreement InterLabs-0015, and the Canada First Research Excellence Fund (CFREF Competition 2, 2015–2016) awarded to the Healthy Brains, Healthy Lives initiative at McGill University, through the Helmholtz International BigBrain Analytics and Learning Laboratory (HIBALL). SBE was supported by the Deutsche Forschungsgemeinschaft (DFG, EI 816/21-1), the National Institute of Mental Health (R01-MH074457), and the European Union’s Horizon 2020 Research and Innovation Programme under Grant Agreement No. 945539 (HBP SGA3).

