## Supplemental file for "Convergent functional effects of antidepressants in major depressive disorder: a neuroimaging meta-analysis"

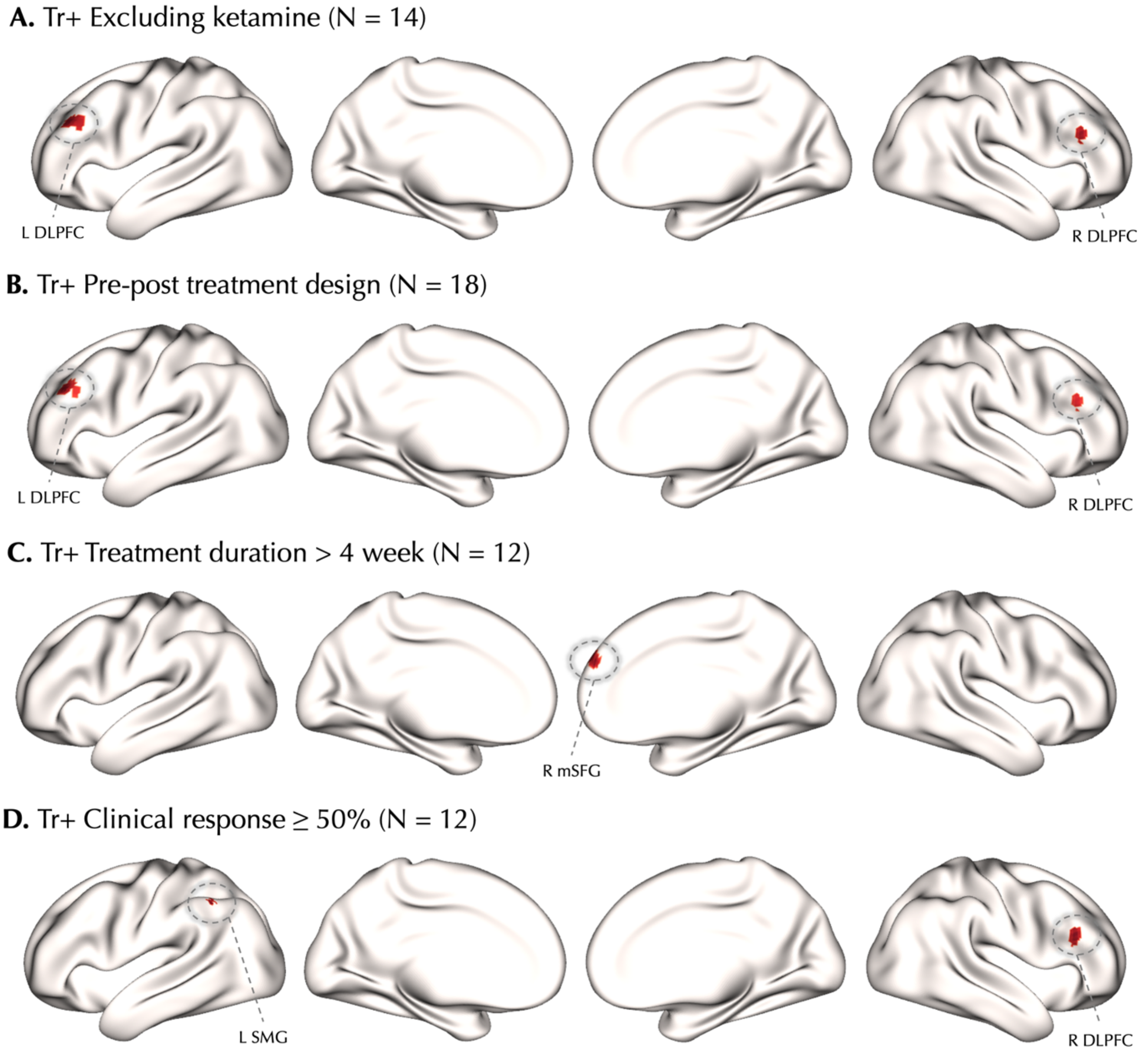


**Fig S1. Subgroup ALE meta-analyses of Treated > Untreated experiments.** The ALE cFWE-corrected significant clusters of convergence in the different subgroups of Treated > Untreated (Tr+) experiments are shown. N represents the number of experiments included in each ALE meta-analysis.

L: left, R: right, DLPFC: dorsolateral prefrontal cortex, mSFG: medial superior frontal gyrus, SMG: supramarginal gyrus

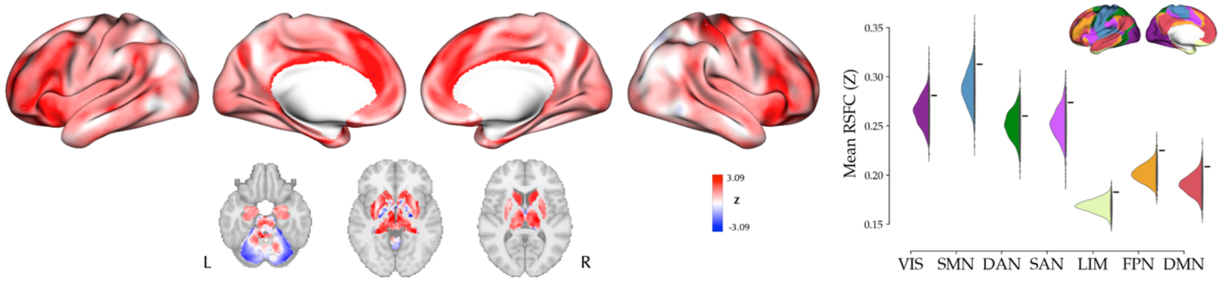


**Fig. S2. Convergent connectivity mapping of antidepressant effects without weighting by sample size.** *left*: The cortical and subcortical map represent Z-scored convergent connectivity map of the foci from all experiments in which the convergent connectivity across experiments is averaged without weighting by their sample sizes. *right*: Mean resting state functional connectivity (RSFC) of the observed foci across canonical resting state networks (denoted by "-") compared against null mean values calculated based on 1000 permutations of randomly selected foci (half-violin plots). None of the networks showed significantly more extreme observed mean RSFC than the null distribution in a two-tailed test and after false discovery rate adjustment at 5%.

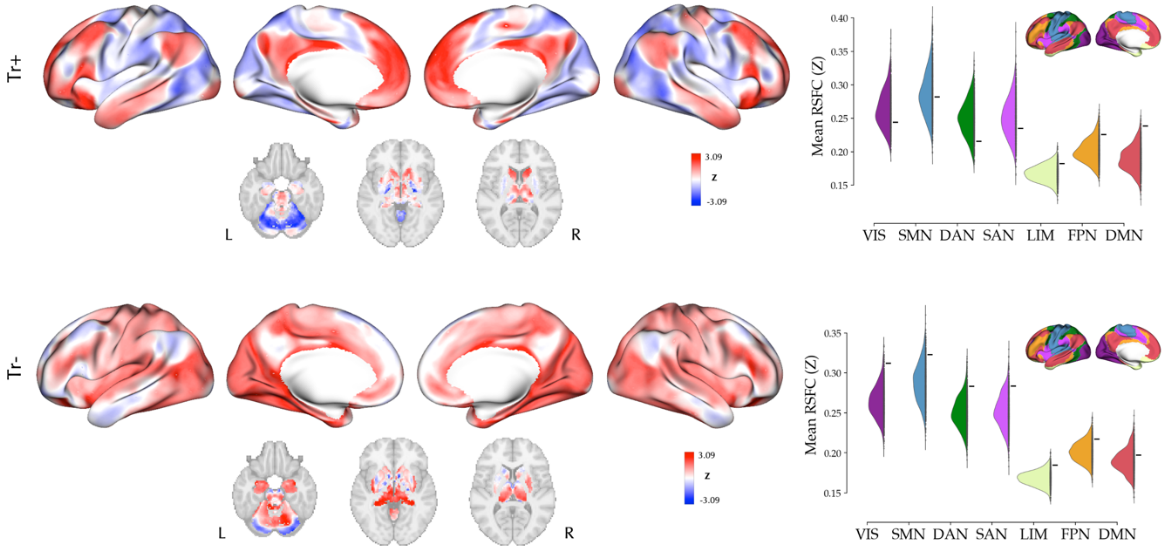


**Fig. S3. Convergent connectivity mapping of antidepressant effects across contrasts.** *left:* The cortical and subcortical maps represent Z-scored convergent connectivity maps of the Treated > Untreated (Tr+; *top*) and Untreated > Treated (Tr-; *bottom*) experiments. *right:* Mean resting state functional connectivity (RSFC) of the observed foci across canonical resting state networks (denoted by "-") compared against null mean values calculated based on 1000 permutations of randomly selected foci (half-violin plots) for the Tr+ (*top*) and Tr- (*bottom*) experiments. In both contrasts none of the networks showed significantly more extreme observed mean RSFC than the null distribution in a two-tailed test and after false discovery rate adjustment at 5%.

VIS: visual network, SMN: somatomotor network, DAN: dorsal attention network, SAN: salience network, LIM: limbic network, FPN: frontoparietal network, DMN: default mode network.

**Table S1.** Search keywords.

| | Search Terms |
|---|---|
| #1 | MDD OR "Major Depressive Disorder" OR "Unipolar Depressive Disorder" OR Depress* |
| #2 | (Antidepres*) OR (SSRI* OR SRI OR SRIs OR "Selective Serotonin Reuptake Inhibitor*" OR "Serotonin Uptake Inhibitor*" OR Dapoxetine OR Citalopram OR Escitalopram OR Fluoxetine OR Fluvoxamine OR Mirtazapine OR Paroxetine OR Sertraline OR Vilazodone OR Zimelidin) OR (SNRI* OR "Serotonin Norepinephrine Uptake Inhibitor*" OR Bicifadine OR Venlafaxin OR Desvenlafaxin OR Duloxetine OR "Duloxetine Hydrochloride" OR Milnacipran OR Levomilnacipran OR Levomepromazine OR Sibutramine OR Bicifadine OR mianserin) OR ("Monoamine Oxidase Inhibitor*" OR MAOI* OR Isocarboxazid OR Phenelizine OR Tranylcypromine OR Selegiline OR Clorgyline OR Moclobemide OR Brofaromine) OR (Tricyclic OR Tetracyclic OR TCA OR TCAs OR Imipramine OR Desipramine OR Trimipramine OR Nortriptyline OR Amitriptyline OR Protriptyline OR Amoxapine OR Doxepin OR Maprotiline OR Clomipramine OR amineptine OR butriptyline OR Demexiptiline OR dibenzepin OR dothiepin OR lofepramine OR melitracen OR noxiptiline OR opipramol OR protriptyline OR quinupramine OR tianeptine OR chlorpoxiten OR amersergide) OR ("NMDA Antagonist*" OR ketamine OR esketamine) OR ("selective noradrenaline reuptake inhibitor*" OR NaRI* OR Reboxetine) OR ("Atypical Antidepres*" OR Mirtazapine OR Nefazodone OR Trazodone OR L-Tryptophan OR agomelatine OR vortioxetine OR Bupropion OR Quetiapine) |
| #3 | fMRI OR "Functional magnetic resonance imaging" OR "functional MRI" OR "resting state" OR "resting-state" OR "gray matter" OR "grey matter" OR "regional cerebral blood flow" OR PET OR "positron emission tomography" OR SPECT OR "Single-photon emission computed tomography" OR ASL OR "arterial spin labeling" OR ALFF OR "Amplitude of low frequency fluctuations" OR fALFF OR "Fractional amplitude of low-frequency fluctuations" OR ReHo OR "Regional Homogeneity" OR ICA OR "Independent Component Analysis" OR Graph |
| | #1 AND #2 AND #3 |

**Table S2.** Methodological details of studies included in the meta-analysis.

| # | First Author, Year [a] | Field strength | Task | Task stimulus | Main Softwares | Contrast | Multiple comparisons correction |
|---|---|---|---|---|---|---|---|
| **1** | Abdallah, C. G., 2017 | 3 T | Rest | - | FSL, AFNI | Pre- vs. post-treatment | 3dClustSim |
| | Murrough, J. W., 2015 | 3 T | Emotion perception | Faces | SPM, AFNI | | |
| **2** | Bremner, J. D., 2007 | - | Emotional word pair retrieval | Words | SPM | Pre- vs. post-treatment | Uncorrected (k > 40, p < 0.005) |
| **3** | Carlson, P. J., 2013 | - | Rest | - | SPM | Pre- vs. post-treatment | RFT |
| **4** | Cheng, Y., 2017 | 1.5 T | Rest | - | SPM, REST | Pre- vs. post-treatment | AlphaSim |
| **5** | Downey, D., 2016 | 3 T | Rest | - | SPM | Time x Group (treated vs placebo MDD) | cFWE |
| **6** | Fonzo, G. A., 2019 | 3 T | Emotional conflict | Faces and words | FSL, SPM, R (fmri package) | Time x Group (treated vs placebo MDD) | FDR |
| **7** | Frodl, T., 2011 | 3 T | Emotion recognition | Faces | SPM | Pre- vs. post-treatment | cFWE |
| **8** | Fu, C. H., 2004, 2007 | 1.5 T | Emotion recognition | Faces | In-house | Time x Group (treated MDD vs HC) | cFWE |
| **9** | Fu, C. H., 2015 | 3 T | Affective facial expressions / Emotional Stroop | Faces; Words | SPM | Pre- vs. post-treatment | cFWE |
| **10** | Gonzalez, S., 2020 | 3 T | Rest | - | FSL, ASLtoolbox | Pre- vs. post-treatment | FDR |
| **11** | Jiang, W., 2012 | 1.5 T | Emotion recognition | Faces | SPM | Pre- vs. post-treatment | Uncorrected |
| **12** | Joe, A.Y., 2006 | - | Rest | - | SPM | Pre- vs. post-treatment | Uncorrected (k > 100 voxels, p < 0.005) |
| **13** | Keedwell, P., 2008 | 1.5 T | Emotion perception | Faces | Institute of Psychiatry | Early vs. late post-treatment | cFWE |
| **14** | Kennedy, S. H., 2001 | - | Rest | - | SPM | Pre- vs. post-treatment | cFWE |
| **15** | Kohn, Y., 2008 | - | Rest | - | SPM | Pre- vs. post-treatment | cFWE |

| | | | | | | | |
|---|---|---|---|---|---|---|---|
| **16** | Komulainen, E., 2018 | 3 T | Emotional word processing | Words | SPM | Treatment vs. placebo | FDR |
| | Komulainen, E., 2021 | 3 T | Listening to emotional narratives | Speech | Intersubject Correlation Toolbox, FSL | Treatment vs. placebo | FDR |
| **17** | Kraus, C., 2019 | 7 T | Cued painful stimulation | Symbols +/- Electrical stimulation | SPM | Treated acute MDD vs. untreated remitted MDD | Uncorrected |
| | Rütgen, M., 2019 | 7 T | Empathy for pain | Faces | SPM, AFNI, FSL | Pre- vs. post-treatment | cFWE |
| **18** | Li, C. T., 2016 | - | Rest | - | SPM | Pre- vs. post-treatment | cFWE |
| **19** | Lopez-Sola, M., 2010 | 1.5 T | Painful stimulation | Auditory tone and heat | SPM | Time x Group (treated MDD vs HC) | Uncorrected |
| **20** | Mayberg, H. S., 2000 | - | Rest | - | In-house | Pre- vs. post-treatment | Uncorrected |
| **21** | Reed, J. L., 2018 | 3 T | Dot probe task with emotional faces | Faces and dots | AFNI | Session (treatment vs. placebo) x Group (MDD vs. HC) | 3dClustSim |
| | Reed, J. L., 2019 | | Emotion recognition | Faces | | | |
| **22** | Robertson, B., 2007 | 1.5 T | Emotional oddball | Pictures and shapes | SPM | Pre- vs. post-treatment | Uncorrected |
| **23** | Sankar, A., 2017 | 3 T | Sternberg (verbal working memory) | Letters | SPM | Pre- vs. post-treatment | cFWE |
| **24** | Sterpenich, V., 2019 | 3 T | Monetary incentive delay task / Emotional judgment | Pictures | SPM | Pre- vs. post-treatment | cFWE |
| **25** | Wagner, G., 2010 | 1.5 T | Stroop Color-Word | Words | SPM | Pre- vs. post-treatment | Uncorrected |
| **26** | Walsh, N. D., 2007 | 1.5 T | N-back | Letters | n.a. | Pre- vs. post-treatment | cFWE |
| **27** | Wang, L., 2014 | 3 T | Rest | - | DPARSF, REST | Pre- vs. post-treatment | AlphaSim |
| | Wang, L., 2017 | | | | | Time x Group (MDD vs. HC) | |
| **28** | Wang, Y., 2012 | 3 T | Emotion recognition | Faces | SPM | Pre- vs. post-treatment | Uncorrected |
| **29** | Williams, R. J., 2021 | 3 T | Emotional face matching | Faces | Sandwich Estimator | Pre- vs. post-treatment | TFCE |

| | | | | | | | |
|---|---|---|---|---|---|---|---|
| | | | | | Toolbox, SPM | | |
| **30** | Yin, Y., 2018 | 3 T | Rest | - | SPM | Pre- vs. post-treatment | AlphaSim |

[a] Publications with overlapping samples are grouped together.

n.a.: not available, FSL: FMRIB Software Library, AFNI: Analysis of Functional NeuroImages, DPARSF: Data Processing Assistant for Resting-State fMRI, REST: REsting State fMRI data analysis Toolkit, cFWE: family-wise error correction at cluster level, FDR: false discovery rate, RFT: random field theory.
